# Early clinical characteristics of Covid-19: scoping review

**DOI:** 10.1101/2020.07.31.20165738

**Authors:** Lakshmi Manoharan, Jonathan W S Cattrall, Carlyn Harris, Katherine Newell, Blake Thomson, Mark G Pritchard, Peter G Bannister, Tom Solomon, Peter W Horby, Louise Sigfrid, Gail Carson, Piero Olliaro

## Abstract

**Background:** The Coronavirus disease 2019 (covid-19) pandemic has spread rapidly across the globe. Accurate clinical characterisation studies conducted early in the pandemic are essential to informing research, diagnosis and clinical management efforts. In this scoping review we identify the clinical characteristics of patients admitted to hospital in the early months of the pandemic, focusing on symptoms, laboratory and imaging findings, and clinical outcomes.

**Methods:** A scoping review. MEDLINE, EMBASE and Global Health databases were searched for studies published from January 1^st^ 2020 to April 28^th^ 2020. Studies which reported on at least 100 hospitalised patients with covid-19 of any age were included.

**Results:** Of 1,249 studies identified through the search 78 studies were eligible for inclusion; one randomized control trial and 77 observational studies presenting data on 77,443 patients admitted with covid-19. Most studies were conducted in China (82%), 9% in the US and 10% in Europe and two studies were set in more than one country. No studies included patients from low and middle income countries. Coagulopathy was underrecognised as a complication in the early months of the pandemic. Use of corticosteroids varied widely, and the use of anticoagulants was reported in only one study. Fever, cough and dyspnoea are less common in older adults; gastrointestinal symptoms, as the only presenting feature was underrecognised. The most common laboratory finding was lymphocytopenia. Inflammatory biomarkers were commonly elevated, including C-reactive protein and interleukin-6. Typical computed tomography findings include bilateral infiltrates however imaging may be normal in early disease. Data on clinical characteristics in children and vulnerable populations were limited.

**Conclusions:** Clinical characterisation studies from early in the pandemic indicated that covid-19 is a multisystem disease, with biomarkers indicating inflammation and coagulopathy. However, early data collection on symptoms and clinical outcomes did not consistently reflect this wide spectrum. Corticosteroid use varied widely, and anticoagulants were rarely used. Clinicians should remain vigilant to the possibility of covid-19 in patients presenting without fever, cough and dyspnoea, particularly in older adults. Further characterisation studies in different at-risk populations is needed.

**Review registration:** Available at https://osf.io/r2ch9

## BACKGROUND

The novel coronavirus (SARS-CoV-2) causing the acute respiratory disease covid-19 has spread globally. As the pandemic has progressed, data have emerged indicating that covid-19 might present without the common symptoms of fever, cough and dyspnoea reported early on during the pandemic and as defined in the WHO clinical case definition[1]. Non-respiratory symptoms have been described increasingly, such as gastrointestinal[2], cardiovascular[3], and neurological symptoms[4]. Biochemical markers and imaging can be important for aiding decision making by identifying patients at risk of severe disease or complications and for optimising patient care. Identifying the spectrum of covid-19 clinical presentations, biochemical markers and imaging findings may assist timely identification of infection and inform clinical management and public health interventions. The aim of this review is to characterise the clinical presentation, biochemical and imaging findings, and clinical outcomes of covid-19 disease from studies published early in the epidemic. The review will also highlight gaps in our knowledge to inform future covid-19 research efforts.

## METHODS

To identify the clinical characteristics of patients admitted to hospital with covid-19 globally, we performed a scoping evidence review, drawing on the PRISMA extension for scoping review guidelines (Figure 1).

**Figure 1:**
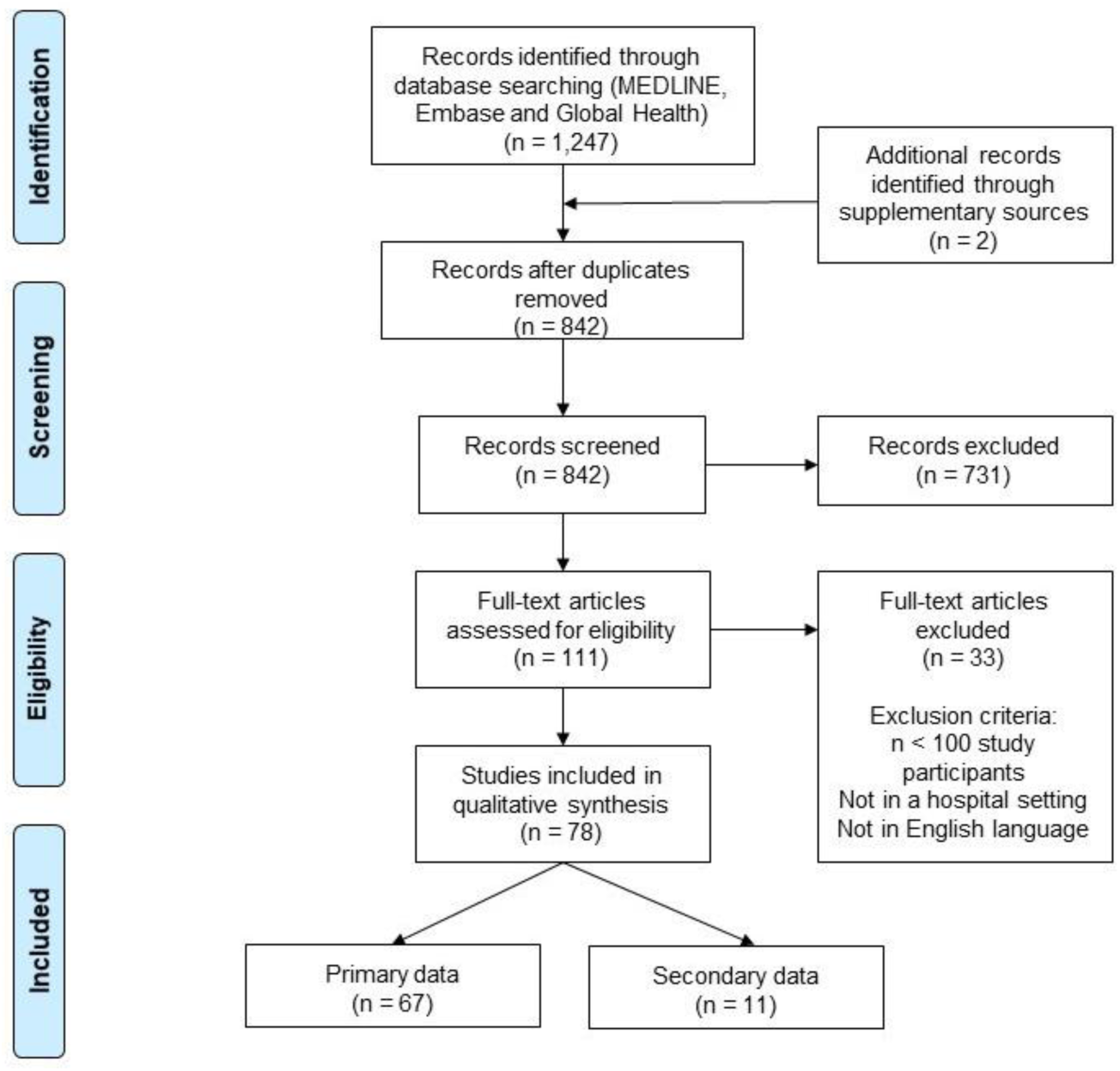
PRISMA diagram for study selection (PRISMA: Preferred Reporting for Systematic Reviews and Meta-Analysis)

### Search strategy

The following databases were searched from inception to 28 April 2020 for relevant studies: MEDLINE; EMBASE and Global Health. The search terms were: (covid-19 OR SARS-CoV-2 OR 2019-nCoV OR novel coronavirus) AND (clinical OR hospital OR admitted) AND (characteristics OR features OR symptoms OR signs), without any further restrictions. The electronic database results were supplemented with a Google Scholar search on the 28 April 2020 with the first 100 results screened for inclusion.

### Screening and eligibility

Two reviewers independently screened the title and abstract of all the retrieved search results, after de-duplication. The full text of the articles that passed the first stage for inclusion were divided and screened by five individual reviewers. Studies presenting clinical data on patients admitted to hospital (ward or ICU) with covid-19 in any hospital globally were included. Due to the number of articles identified studies which presented fewer than 100 covid-19 cases were excluded to ensure comprehensive data. Articles not published in English were excluded due to constraints in reliably translating clinical data.

### Data synthesis

A standardised form for data extraction was developed and piloted by the reviewers before being finalised. A second reviewer independently checked the extracted data. The data extraction and synthesis were performed using Microsoft Excel. Data on study source, month of publication, period when cases were collected, patient demographic, clinical signs and symptoms on admission, biochemical laboratory and imaging results and outcomes were extracted by one reviewer. A second reviewer independently checked the extracted data. The tabulated data were analysed and synthesised by all reviewers.

## RESULTS

A total of 77 observational studies and one randomised controlled trial (RCT) presenting clinical data on 77,443 patients admitted to hospital with covid-19 in seven countries were included and presented in this review (Figure 1). The majority of studies were from China (82%), followed by the USA (9%), Italy (4%), France (3%) and the UK (3%). China, the USA and the UK contributed 39%, 36% and 22% of patients, respectively. Two studies were set in more than one country, with one including patients from both the US and China, (n =424 patients) and one study including patients from Belgium, France, Italy and Spain (n=417 patients). (Table 1 and Figure 2). There were no studies including samples from populations in low and middle income countries (LMICs). The earliest data of admitted patients were collected from the 21^st^ November 2019 and the latest cohort of data were collected up until18^th^ April 2020.

**Table 1.** Number of studies by country and covid-19 patients included

|  | Number of studies | Total number of patients |
| --- | --- | --- |
| **China** | 63 | 30,082 |
| **USA** | 6 | 27,500 |
| **Italy** | 3 | 1,932 |
| **UK** | 2 | 16,850 |
| **France** | 2 | 238 |
| **USA/China†** | 1 | 424 |
| **Belgium/France/Italy/Spain\*** | 1 | 417 |
| **Total:** | 78 | 77,443 |

†n patients from China = 219, n patients from USA = 205
\*n of patients from each country not delineated

**Figure 2.**
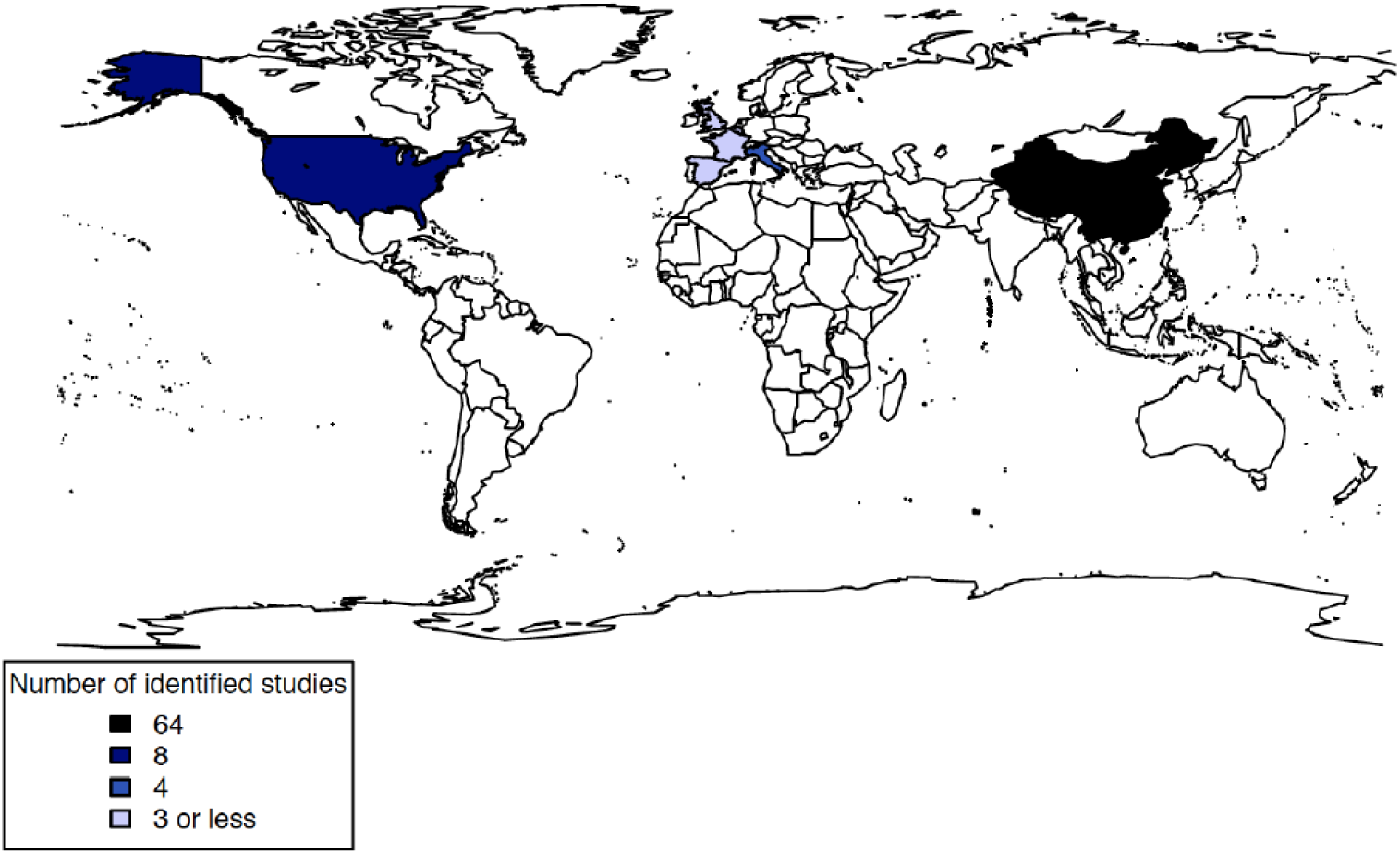
Geographical coverage of the included studies

In the included studies, 68% (n=53) presented clinical data on adults only, 1% (n=1) on children only (<18 years), 23% (n=18) on both adults and children, 1% (n=1) on older adults and 1% (n=1) on pregnant women. A summary table of all included studies is available. (See Additional file 1).

### Clinical presentation

The clinical presentation of covid-19 includes a wide spectrum of disease manifestations. The most commonly reported symptom in the included studies was fever, reported in 77% of articles, followed by cough in 71% of articles. These symptoms, both part of the clinical case definition of covid-19, had a wide prevalence range reported across studies. The highest prevalence of fever (98.6%) was reported in a study of 138 patients with a median age of 56[5] and the highest prevalence of cough (87%) was reported in a study of 114 patients with a mean age of 47.[6] The lowest prevalence of both fever (8.9%) and cough (4%) was reported in a study by Lovell et al. of 101 palliative patients with a median age of 81.^6^ Dyspnoea was the third most reported symptom, included in 62% of all studies. The highest reported prevalence of dyspnoea (80%) was in a study of 178 patients, of which 75% were 50 years or older.[7] The lowest reported prevalence of dyspnoea (0.8%) was in a retrospective study of 118 patients with mean age of 44 years.

Other commonly reported symptoms included fatigue, reported in 47% of articles, myalgia in 42%, sore throat in 40%, chest pain in 25%, rhinorrhoea in 22%, and expectoration in 13%. Noticeably, many patients develop gastrointestinal symptoms including diarrhoea which was reported in 58% of articles, nausea and/or vomiting (40%) and appetite changes (16%).

Gastrointestinal symptoms in the absence of respiratory symptoms were reported in 4% of patients on admission in a cohort of 16,749 covid-19 patients in the UK.[8] Neurological symptoms were also reported in patients, including headache in 44% of articles, altered mental state in 10%, and smell or taste disturbances in 4%. The most commonly reported symptoms and the studies reporting the highest and lowest prevalence of each are reported below (Table 2).

**Table 2.** Prevalence of reported covid-19 symptoms. The table shows the most common symptoms reported and the studies reporting the lowest and highest prevalence of each.

| Clinical syndrome | Presenting symptom/sign | N (%) of all studies | Lowest prevalence reported |  |  | Highest prevalence reported |  |  |
| --- | --- | --- | --- | --- | --- | --- | --- | --- |
|  |  |  | %<sup>ref</sup> | Sample size | Age range | %<sup>ref</sup> | Sample size | Age range |
| Respiratory | Cough (dry/productive) | 55 (71.4) | 4.0 [15] | 101 | 82(72-89)<sup>+</sup> | 87.0 [6] | 114 | 47(16)<sup>*</sup> |
|  | Dyspnoea | 48 (62.3) | 0.8 [16] | 118 | 44.1(13.6)<sup>*</sup> | 80.0 [7] | 178 | ≥50<sup>□</sup> |
|  | Sore throat | 31 (40.3) | 0.7 [17] | 298 | 47(33-61)<sup>+</sup> | 43.0 [6] | 114 | 47(16)<sup>*</sup> |
|  | Chest pain | 19 (24.7) | 1.6 [18] | 125 | 38.8(13.8)<sup>*</sup> | 65.2 [19] | 112 | 65(49-70.8)<sup>+</sup> |
|  | Expectoration | 19 (13.0) | 4.4 [20] | 137 | 57<sup>+</sup> | 34.9 [21] | 645 | 46.7(13.8)<sup>*</sup> |
|  | Rhinorrhoea | 17 (22.1) | 1.0 [17] | 298 | 47(33-61)<sup>+</sup> | 35.3 [22] | 136 | 69(61-77)<sup>+</sup> |
| Gastrointestinal | Diarrhoea | 45 (58.4) | 1.0 [23] | 131 | 47(15)<sup>*</sup> | 52.0 [6] | 114 | 47(16)<sup>*</sup> |
|  | Nausea and/or vomiting | 31 (40.3) | 0.5 [24] | 889 | 47.8(15.2)<sup>*</sup> | 35.0 [6] | 114 | 47(16)<sup>*</sup> |
|  | Changes in appetite and or /anorexia | 12 (15.6) | 3.0 [25] | 120 | 45.4(15.6)<sup>*</sup> | 78.6 [26] | 204 | 52.9(16)<sup>*</sup> |
| Neurological | Headache | 34 (44.2) | 0.9 [27] | 108 | 52(37-58)<sup>+</sup> | 82.0 [6] | 114 | 47(16)<sup>*</sup> |
|  | Altered mental state (confusion/agitation) | 8 (10.4) | 1.4 [28] | 1590 | 44.2(14.8)<sup>*</sup> | 42.6 [15] | 101 | 82(72-89)<sup>+</sup> |
|  | Smell or taste impairment | 3 (3.9) | 5.6 [9] | 214 | 52.7(15.5)<sup>*</sup> | 88.8 [29] | 417 | 36.9(11.4)<sup>+</sup> |
| Systemic | Fever | 59 (76.6) | 8.9 [15] | 101 | 82(72-89)<sup>+</sup> | 98.6 [5] | 138 | 56(42-69)<sup>+</sup> |
|  | Myalgia and/or arthralgia | 32 (41.6) | 2.0 [23] | 131 | 47(15)<sup>*</sup> | 74.0 [6] | 114 | 47(16)<sup>*</sup> |
|  | Fatigue | 36 (46.8) | 4.7 [17] | 298 | 47(33-61)<sup>+</sup> | 93.0 [6] | 114 | 47(16)<sup>*</sup> |

+ Median (IQR), \*Mean (SD), <sup>□</sup>75% of patients aged ≥ 50

In addition, covid-19 complications and their associated symptoms may be the presenting features of the disease. In a study of 214 covid-19 infected patients, acute cerebrovascular events affected 2.8% of included participants, with two of these patients presenting to the Emergency Department with neurological symptoms in the absence of respiratory symptoms.[9] Five of the included studies reported data on covid-19 incubation period, with the minimum and maximum incubation period reported in studies ranging from 3 to 6.7 days.[10-14]

### Laboratory findings

Laboratory results were reported in 54 studies. Although most studies reported laboratory values on admission, collection timepoint was not always defined. Lymphocytopenia was the most common laboratory finding among patients admitted to hospital with confirmed covid-19, being reported in 32% (25/78) of studies[11, 21, 30-35]. The highest prevalence of lymphocytopenia (99.1%) was reported in a study of 225 patients with a mean age of 50 years[36]. The second most commonly reported laboratory finding was elevated C-reactive protein (CRP) in 19% (15/78) of studies. The highest prevalence of elevated CRP (91.9%) was reported in a study of 140 patients with a mean age of 57 years[37].

Other abnormal laboratory findings included thrombocytopenia, elevated erythrocyte sedimentation rate (ESR), lactate dehydrogenase (LDH), D-dimer, troponin and cytokine levels, particularly interleukin 6 (IL-6). Aspartate aminotransferase (AST) and alanine aminotransferase (ALT) were both elevated, and decreased albumin and leukopenia were reported in several studies[11, 21, 30-35, 38-41]. (Table 3)

**Table 3.**
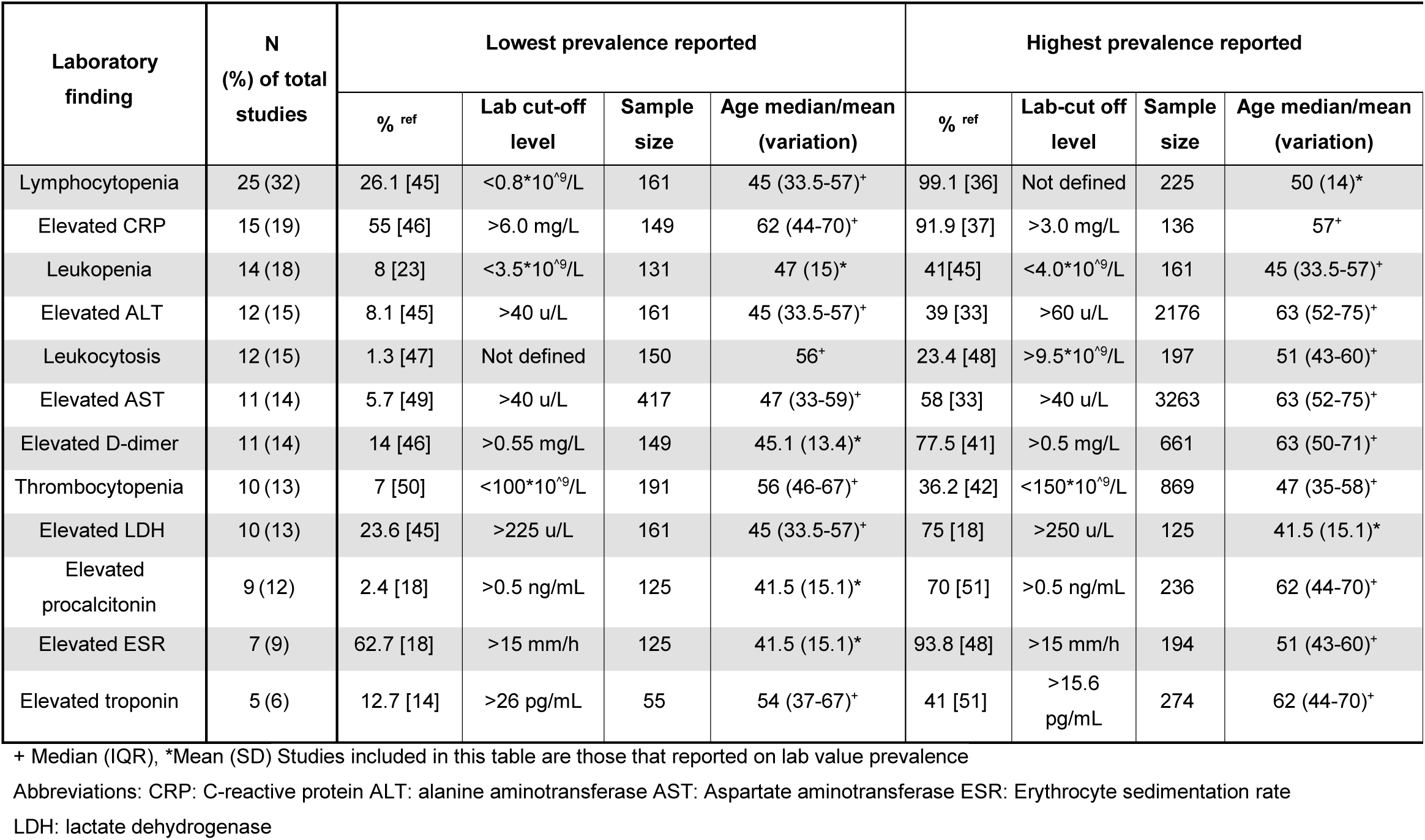
Abnormal laboratory findings in covid-19. The table shows the studies that reported the lowest and highest prevalence of the most common abnormal laboratory findings reported, as assessed by the study authors.

The largest study identified which reported laboratory results presented data on 5700 adults and children with laboratory confirmed SARS-CoV-2 infection admitted to hospitals in New York. Most of these results were collected within 24 hours of admission.[33] This study reported that 60% (n=3387) of patients presented with lymphocytopenia, 58% (n=3263) had elevated AST, 39% (n=2176) elevated ALT, 22.6% (n=801) and an elevated troponin I or troponin T. This study also showed evidence of elevated BNP (median = 385 pg/mL, n= 1818), D-dimer (median= 438 ng/mL, n= 3169), procalcitonin (median= 0.2ng/mL, n=4138), LDH (median= 404 U/L, n=4003), and ferritin (median= 798 ng/mL, n=4344)[33].

In studies which compared laboratory values among mild and severe cases of covid-19, those with severe disease were found to have more prominent lab abnormalities including lower lymphocyte counts, higher inflammatory marker levels, (CRP, ESR, LDH), and elevated D-dimer levels and liver enzymes (AST, ALT) [17, 31, 34, 35, 42-44]. Among studies that reported immune markers, higher levels of serum cytokines and lower levels of T lymphocytes were associated with disease severity[35, 43].

### Imaging findings

Imaging findings were reported in 44 studies. Common abnormalities seen on computed tomography (CT) at admission included ground glass opacities and consolidation (Table 4) [5, 31, 34, 36, 50-52]. These lesions were predominantly bilateral[31, 34, 48, 53, 54] and frequently affected the lower lobes[21, 47, 54, 55]. Reported prevalence of ground glass opacities ranged from 12.1% to 96% in the included studies[31, 46]. Other frequently reported covid-19 CT imaging features include a peripheral distribution of lesions[16, 25, 46, 56, 57], and multi-lobar involvement[23, 35, 54, 55]. The prevalence of lesions in two or more lobes ranged from 35.8% to 94.6%[21, 35]. Less common CT abnormalities include pleural effusion and lymphadenopathy[25, 31, 58]. Abnormal CT findings were more prevalent in those with severe disease[17, 31, 35, 40].

**Table 4.**
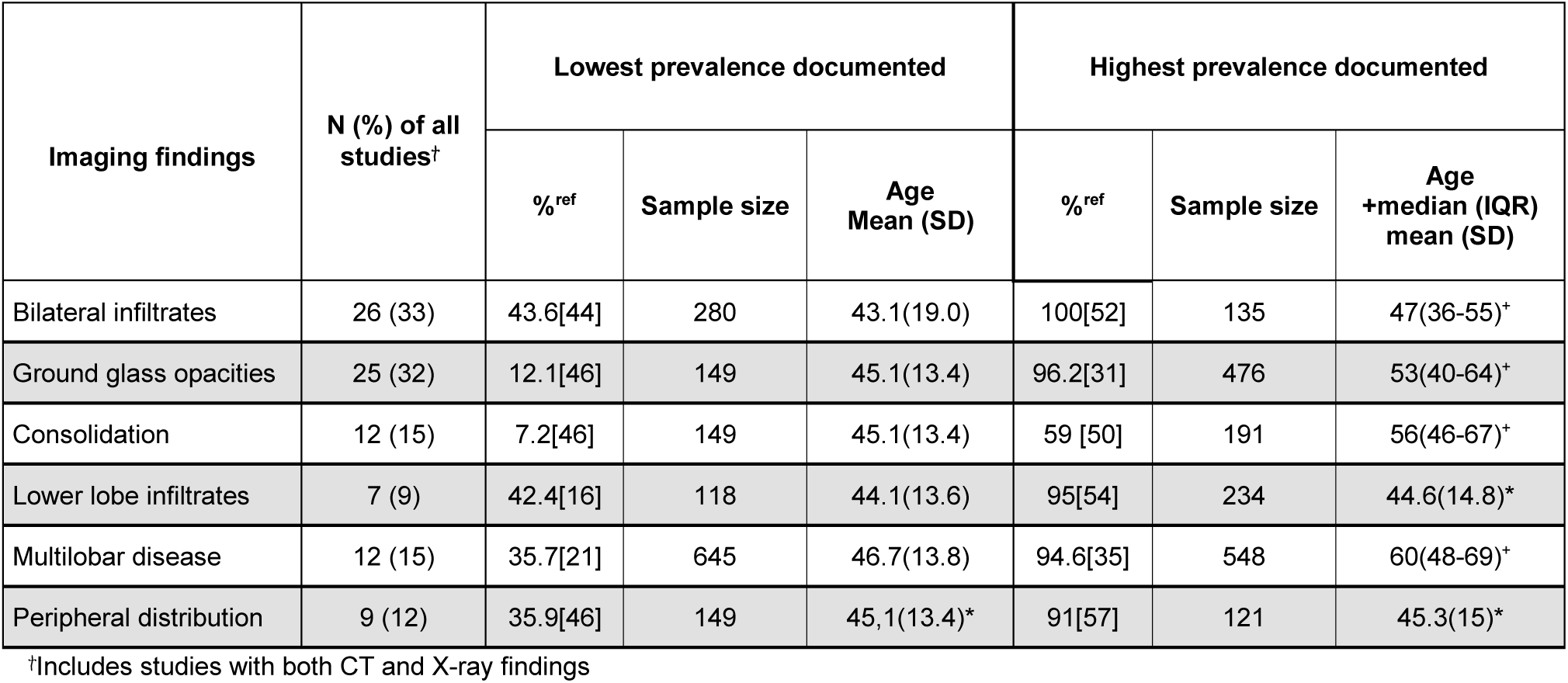
Imaging findings in covid-19. The table shows the most common imaging findings reported and the studies reporting the lowest and highest prevalence of each.

The included studies demonstrated that imaging findings may be normal in early or mild disease. In a study of 298 laboratory confirmed patients, 14.8% (44/298) of all patients had a normal chest CT on admission[17]. CT imaging was more likely to be normal the sooner it was conducted after symptom onset[17]. In a study of 121 laboratory confirmed patients, 56% (n=20/36) of patients had a normal CT scan 0 to 2 days post-symptom onset[57], while a study of 112 patients reported a normal scan in 21% (n=10/47) of patients 0 to 4 days post-symptom onset[59]. A study of 543 patients admitted to a Chinese hospital found that the median time from symptom onset to the diagnosis of pneumonia on CT was 4 days[35]. Disease progression on repeated imaging was reported in a small number of studies. In a study of 248 confirmed cases who had repeated radiological examination, 66% (n=163/248) showed disease progression after a median of 3 days[60], while in a study of 149 patients where 17 had initial normal radiological findings, 29% (5/17) had disease progression on imaging after a median of 7 days[46].

There were 10 studies which reported both CT and chest X-ray findings[17, 21, 27, 28, 42, 44, 48, 50, 58, 60]. The prevalence of abnormalities was greater with CT imaging in studies that reported findings of both modalities. In a cohort of 1,590 laboratory confirmed patients from multiple hospitals in China, 15% (n=243/1590) of chest X-ray findings on admission were abnormal compared to 71% (n=1130/1590) of CT results.[28] In a study of 1099 laboratory confirmed patients, 59% (n=162/274) of those who had a chest X-ray on admission demonstrated abnormalities compared to 86% (n=840/975) of those who had a CT[42]. Other studies which reported both CT and X-ray imaging did not individually present results, instead combining the two modalities when reporting imaging abnormalities[17, 21, 44, 48, 50, 60].

### Outcomes

Mortality rate among hospitalised cases was reported to be up to 33%[8], with higher mortality rates observed in studies of ICU patients ranging from 45%[8] to 50%.[61] Commonly documented complications and adverse outcomes included acute respiratory distress syndrome (ARDS) [14, 48, 50, 60], ICU treatment[2, 6, 11, 14, 19, 28, 33, 41, 42, 48, 50, 55, 60, 62, 63], mechanical ventilation[11, 28, 33, 41], and death[2, 5, 6, 8, 11, 12, 14, 15, 17-20, 25-27, 30-36, 38-41, 44, 46, 48, 50-53, 55, 60-69]. Several studies reported that older patients were more prone to serious adverse outcomes, with age a documented risk factor for ARDS[48], ICU admission[60] and in-hospital mortality[8, 14, 30, 31, 34, 51, 64].

Males were reported as more susceptible to these adverse outcomes, and in a study of 1591 laboratory confirmed ICU patients set in Italy, 82% (n=1304/1591) were male[64]. This was corroborated in another four studies reporting higher mortality rates and disease severity for men amongst critically[8, 33, 51] and non-critically ill patients[40], including in a study of 16,747 patients from the UK which found that female sex was significantly associated with a reduced risk of mortality (hazard ratio 0.80 [95% CI 0.72, 0.89]) compared to male[8]. A large multi-site study of 5700 patients in New York did not document any deaths in patients under 20 years[33].

Reporting of haematological complications was limited, and only one study from China reported coagulopathy observed in 19% (n=37/191) of patients[50]. Renal complications were reported, with Cheng et al.[41] documenting acute kidney injury (AKI) in 5.1% (n=36/701) of patients. In this study, AKI was found to be an independent predictor of in-hospital death[41]. AKI was also documented as a complication in 15% (n=28/191) of 191 hospitalised patients.[50] Cardiovascular complications were also reported, and in a study of 393 patients, arrythmias and myocardial infarction was reported in 7.4% and 3.6% of patients, respectively[39]. In a European study including patients from four countries, olfactory dysfunction was prevalent in 63% of patients with clinically resolved infection[29].

Corticosteroid use was higher in patients with severe disease[11, 17, 31, 53]. Rates of corticosteroid use ranged up to 73%[38]. Respiratory support status in patients receiving corticosteroids was not reported in any study. A study of 548 patients reported that high dose corticosteroids was associated with increased risk of death[35], while several other studies suggested high doses were associated with poor clinical outcomes[31, 50, 67]. Most studies did not recommend the use of corticosteroids due to absence of reliable evidence[18, 51, 60]. Anticoagulant administration was reported in only one study of 449 patients[65]. This study demonstrated that in those patients with elevated D-dimer, there was a significantly lower mortality rate in those patients administered heparin compared to those who were not.

## DISCUSSION

In this scoping review we present clinical characteristics of more than 77,000 people admitted to hospital with covid-19 in seven countries, focusing on symptoms, laboratory and imaging findings and clinical outcomes.

Although fever and cough were the most commonly reported symptoms they may not be present all the time in patients across all age-ranges. For instance, in the study by Lovell et al. of 101 elderly palliative care patients, less than ten percent presented with fever or cough[15]. This is also consistent with the International Severe Acute Respiratory and emerging Infection Consortium (ISARIC) database which demonstrates a lower proportion of older adults presented with fever, cough and dyspnoea compared to younger adults[70].

These data indicate that the covid-19 clinical syndrome may vary with age, with older adults presenting differently and that further clinical characterisation is needed in this age group. Furthermore, these data have shown that patients may present with non-respiratory symptoms, as described in the ISARIC report where 4% of patients presented with gastrointestinal symptoms alone[8]. The findings show relying solely on respiratory symptoms would potentially misdiagnose the condition particularly in children/adolescents and in elderly patients.

Laboratory and imaging findings are also diverse, and although CT is more sensitive in detecting lung abnormalities it can also be normal early on in the disease[17, 57, 59]. Although lymphocytopenia and ground glass opacities on CT were the most frequently reported laboratory and radiological findings, these abnormalities may be absent at presentation and therefore normal findings alone cannot rule out covid-19. The wide variation in prevalence of abnormal findings in hospitalised patients may be partly due to the difference in populations included and timing of sampling, but also shows emerging evidence of multiple organs being affected. These biochemical data indicate increased risk of inflammatory and thromboembolic disease, and are in line with suggestions that SARS-CoV-2 induces ARDS and overproduction of proinflammatory cytokines. Immunomodulatory drugs may be beneficial through controlling inflammation, but their use must be balanced against the theoretical risk of allowing uncontrolled pathogen replication[71].

One study focussed solely on evaluating the clinical characteristics of children[72], and whilst some studies included patients of all ages, these had low numbers of children and wide age brackets. The paucity of evidence regarding children likely reflects a reduced need for hospitalisation due to lower clinical severity of covid-19, but also indicates a need for more paediatric studies. It is important to investigate the full range of potential clinical presentations in children, especially given increasing case reports of severe multisystem inflammatory conditions in children[73]. Nearly 80% of included studies and more than one-third of patients were from China, and ethnicity was not reported in these studies. As increasing evidence emerges regarding the complex relationship between ethnicities and other risk factors for covid-19 severity[74, 75], this relationship will be important to elicit in future studies in different settings.

The studies in this review collected data on patients admitted to hospital in the early months of the pandemic, when awareness of non-respiratory symptoms was lower and therefore may not have been recognised or documented by healthcare providers or researchers. Anosmia and dysgeusia have been reported increasingly[76] and has been included in several countries’ covid-19 case definition[77-79]. Coagulation abnormalities were not frequently reported in these studies. However, there is increasing evidence regarding the higher risk of thrombosis in covid-19 patients suggesting that monitoring of coagulopathy and use of anti-thrombotic medication may be an important aspect of treatment[80, 81]. Use of anticoagulants was reported in only one study[65]. Other manifestations emerging from recent literature include neurological[4] and cardiovascular[3] disease. Although biochemical parameters from early studies in China indicated multiorgan manifestations, these outcomes were not frequently reported in the included studies[50]. The long term outcomes of covid-19 are still unknown, however patients with severe disease may have significant complications. As our understanding of the type and frequency of covid-19 symptoms evolves, this should be reflected in updated patient data collection protocols to ensure emerging symptoms are documented[82].

The use of corticosteroids varied widely in the included studies with administration of different dose, duration and target populations, and respiratory support status was not provided for these patients. Although most of studies from early in the pandemic recommended against the use of corticosteroids, recent evidence from the RECOVERY RCT demonstrates a reduction in mortality with use of low dose dexamethasone in patients requiring respiratory support. This highlights the importance of integrating evidence from RCTs into clinical management and national health authority guidelines during a pandemic.

As the covid-19 pandemic has progressed across geographical boundaries and populations, there is increasing data available from other countries and specifically from LMICs. Our review included studies published before the 28^th^ April, and we found no studies reporting patient cohorts from LMICs despite both Egypt and Brazil reporting initial covid-19 cases in February 2020. This may be due to under-recognition or underreporting of cases in these countries earlier on in the pandemic.

There were limitations to this review, we focused on hospitalised patients due to limited data being available from primary care settings. We only included studies with at least 100 patients to ensure robustness. Hospitalised patients are likely to represent the more severe end of the clinical spectrum, presenting with a more advanced clinical picture compared to cases in the community. As reports emerge of the elderly with severe disease being cared for in care homes, this population may be under-represented in our data. New symptoms that have recently become significant may not have been documented consistently, and the data biased by different countries’ admission policies. Furthermore, evaluating the progression of clinical characteristics was challenging as many studies did not report on the day of illness on which results were recorded.

## CONCLUSION

This review reflects our knowledge of covid-19 in the earlier months of the pandemic. Awareness of presentation with non-respiratory symptoms has increased over time, which has significant implications for the identification of cases and prevention of nosocomial and care home transmission. These factors are likely important amplifiers of local outbreaks and disproportionately affect vulnerable populations. These data highlight the urgent need for studies to include different at risk populations and emphasise the importance of clinical characterisation studies which facilitate robust and comparable data collection.

## Data Availability

Data sharing is not applicable to this article as no datasets were generated or analysed during the current study.

## DECLARATIONS

### Ethics approval and consent to participate

Not applicable.

### Consent for publication

Dissemination to participants and related patient and public communities: No patients were directly involved in this review. The research findings will be disseminated using the network of international researchers, ISARIC, which comprises over 100 international groups studying epidemic readiness. Additionally, it will be disseminated through ALERRT, a group of Africa specific research groups. The research will be displayed on the Epidemic diseases Research Group Oxford (ERGO) website and through its social media service.

### Competing interests

The authors declare that they have no competing interests.

### Funding

This work was funded by the Department of Health and Social Care using UK Aid funding and is managed by the NIHR. The views expressed in this publication are those of the author(s) and not necessarily those of the Department of Health and Social Care.

This work was supported by the Wellcome Trust. The funder has no role in study design, data collection, data analysis or writing of the report.

TS is supported by the National Institute for Health Research (NIHR) Health Protection Research Unit in Emerging and Zoonotic Infections (Grant No. IS-HPU-1112-10117 and NIHR200907). NIHR Programme Grant for Applied Research (No. RP-PG-0108-10,048), NIHR Global Health Research Group on Brain Infections (No. 17/63/110), and the European Union’s Horizon 2020 research and innovation program ZikaPLAN (Preparedness Latin America Network), grant agreement No. 734584.

### Authors’ contributions

GC and PO conceived the project. BT and LM led the project, overseeing screening and data extraction and writing the manuscript. BT performed the literature search. LM, JC, CH, KN, BT, PB and MP screened the references and extracted data. LM, JC, CH, KN, PB and LS contributed to the writing of the manuscript. LS provided initial feedback and edits. PO, GC, PH, TS and LS provided comments and feedback on the project. PO is the group leader and provided leadership on data interpretation. The corresponding author attests that all listed authors meet authorship criteria and that no others meeting the criteria have been omitted.

## Notes

### Competing Interest Statement

The authors have declared no competing interest.

### Clinical Protocols

https://osf.io/r2ch9

### Author Declarations

Ethics committee approval not required for this scoping review using published studies

## References

1. Global surveillance for COVID-19 caused by human infection with COVID-19 virus. World Health Organisation (WHO); 2020 18th June 2020.

2. Team CC-R. Characteristics of Health Care Personnel with COVID-19 -United States, February 12-April 9, 2020. MMWR Morb Mortal Wkly Rep. 2020;69(15):477–81.

3. Bonow RO, Fonarow GC, O’Gara PT, Yancy CW. Association of Coronavirus Disease 2019 (COVID-19) With Myocardial Injury and Mortality. JAMA Cardiology. 2020.

4. Ellul M, Benjamin L, Singh B, Lant S, Michael B, Easton A, et al. Neurological Associations of COVID-19. The Lancet Neurology – IN PRESS June 2020. 2020.

5. Wang D, Hu B, Hu C, Zhu F, Liu X, Zhang J, et al. Clinical Characteristics of 138 Hospitalized Patients With 2019 Novel Coronavirus-Infected Pneumonia in Wuhan, China. JAMA. 2020.

6. Klopfenstein T, Kadiane-Oussou NJ, Toko L, Royer PY, Lepiller Q, Gendrin V, et al. Features of anosmia in COVID-19. Med Mal Infect. 2020.

7. Garg S, Kim L, Whitaker M, O’Halloran A, Cummings C, Holstein R, et al. Hospitalization Rates and Characteristics of Patients Hospitalized with Laboratory-Confirmed Coronavirus Disease 2019 - COVID-NET, 14 States, March 1-30, 2020. MMWR Morb Mortal Wkly Rep. 2020;69(15):458–64.

8. Docherty AB, Harrison EM, Green CA, Hardwick HE, Pius R, Norman L, et al. Features of 16,749 hospitalised UK patients with COVID-19 using the ISARIC WHO Clinical Characterisation Protocol. medRxiv. 2020:2020.04.23.20076042.

9. Mao L, Jin H, Wang M, Hu Y, Chen S, He Q, et al. Neurologic Manifestations of Hospitalized Patients With Coronavirus Disease 2019 in Wuhan, China. JAMA Neurol. 2020.

10. Zhang J, Litvinova M, Wang W, Wang Y, Deng X, Chen X, et al. Evolving epidemiology and transmission dynamics of coronavirus disease 2019 outside Hubei province, China: a descriptive and modelling study. Lancet Infect Dis. 2020.

11. Guan W-J, Ni Z-Y, Hu Y, Liang W-H, Ou C-Q, He J-X, et al. Clinical Characteristics of Coronavirus Disease 2019 in China. The New England journal of medicine. 2020;382(18):1708–20.

12. Tian S, Hu N, Lou J, Chen K, Kang X, Xiang Z, et al. Characteristics of COVID-19 infection in Beijing. J Infect. 2020;80(4):401–6.

13. Lian J, Jin X, Hao S, Cai H, Zhang S, Zheng L, et al. Analysis of Epidemiological and Clinical features in older patients with Corona Virus Disease 2019 (COVID-19) out of Wuhan. Clin Infect Dis. 2020.

14. Cao J, Tu WJ, Cheng W, Yu L, Liu YK, Hu X, et al. Clinical Features and Short-term Outcomes of 102 Patients with Corona Virus Disease 2019 in Wuhan, China. Clin Infect Dis. 2020.

15. Lovell N, Maddocks M, Etkind SN, Taylor K, Carey I, Vora V, et al. Characteristics, symptom management and outcomes of 101 patients with COVID-19 referred for hospital palliative care. J Pain Symptom Manage. 2020.

16. Zhao W, Zhong Z, Xie X, Yu Q, Liu J. CT Scans of Patients with 2019 Novel Coronavirus (COVID-19) Pneumonia. Theranostics. 2020;10(10):4606–13.

17. Cai Q, Huang D, Ou P, Yu H, Zhu Z, Xia Z, et al. COVID-19 in a designated infectious diseases hospital outside Hubei Province, China. Allergy. 2020.

18. Wang R, Pan M, Zhang X, Fan X, Han M, Zhao F, et al. Epidemiological and clinical features of 125 Hospitalized Patients with COVID-19 in Fuyang, Anhui, China. Int J Infect Dis. 2020.

19. Deng Q, Hu B, Zhang Y, Wang H, Zhou X, Hu W, et al. Suspected myocardial injury in patients with COVID-19: Evidence from front-line clinical observation in Wuhan, China. Int J Cardiol. 2020.

20. Liu K, Fang YY, Deng Y, Liu W, Wang MF, Ma JP, et al. Clinical characteristics of novel coronavirus cases in tertiary hospitals in Hubei Province. Chin Med J (Engl). 2020;133(9):1025–31.

21. Zhang X, Cai H, Hu J, Lian J, Gu J, Zhang S, et al. Epidemiological, clinical characteristics of cases of SARS-CoV-2 infection with abnormal imaging findings. Int J Infect Dis. 2020;94:81–7.

22. Shao F, Xu S, Ma X, Xu Z, Lyu J, Ng M, et al. In-hospital cardiac arrest outcomes among patients with COVID-19 pneumonia in Wuhan, China. Resuscitation. 2020;151:18–23.

23. Li X, Zeng W, Li X, Chen H, Shi L, Li X, et al. CT imaging changes of corona virus disease 2019(COVID-19): a multi-center study in Southwest China. J Transl Med. 2020;18(1):154.

24. Wang G, Chen W, Jin X, Chen YP. Description of COVID-19 cases along with the measures taken on prevention and control in Zhejiang, China. J Med Virol. 2020.

25. Zhang R, Ouyang H, Fu L, Wang S, Han J, Huang K, et al. CT features of SARS-CoV-2 pneumonia according to clinical presentation: a retrospective analysis of 120 consecutive patients from Wuhan city. Eur Radiol. 2020.

26. Pan L, Mu M, Yang P, Sun Y, Wang R, Yan J, et al. Clinical Characteristics of COVID-19 Patients With Digestive Symptoms in Hubei, China: A Descriptive, Cross-Sectional, Multicenter Study. Am J Gastroenterol. 2020;115(5):766–73.

27. Yao Q, Wang P, Wang X, Qie G, Meng M, Tong X, et al. Retrospective study of risk factors for severe SARS-Cov-2 infections in hospitalized adult patients. Pol Arch Intern Med. 2020.

28. Liang WH, Guan WJ, Li CC, Li YM, Liang HR, Zhao Y, et al. Clinical characteristics and outcomes of hospitalised patients with COVID-19 treated in Hubei (epicenter) and outside Hubei (non-epicenter): A Nationwide Analysis of China. Eur Respir J. 2020.

29. Lechien JR, Chiesa-Estomba CM, De Siati DR, Horoi M, Le Bon SD, Rodriguez A, et al. Olfactory and gustatory dysfunctions as a clinical presentation of mild-to-moderate forms of the coronavirus disease (COVID-19): a multicenter European study. Eur Arch Otorhinolaryngol. 2020.

30. Wang L, He W, Yu X, Hu D, Bao M, Liu H, et al. Coronavirus disease 2019 in elderly patients: Characteristics and prognostic factors based on 4-week follow-up. J Infect. 2020.

31. Feng Y, Ling Y, Bai T, Xie Y, Huang J, Li J, et al. COVID-19 with Different Severity: A Multi-center Study of Clinical Features. Am J Respir Crit Care Med. 2020.

32. Zhang JJ, Cao YY, Dong X, Wang BC, Liao MY, Lin J, et al. Distinct characteristics of COVID-19 patients with initial rRT-PCR-positive and rRT-PCR-negative results for SARS-CoV-2. Allergy. 2020.

33. Richardson S, Hirsch JS, Narasimhan M, Crawford JM, McGinn T, Davidson KW, et al. Presenting Characteristics, Comorbidities, and Outcomes Among 5700 Patients Hospitalized With COVID-19 in the New York City Area. JAMA. 2020.

34. Zhang J, Wang X, Jia X, Li J, Hu K, Chen G, et al. Risk factors for disease severity, unimprovement, and mortality in COVID-19 patients in Wuhan, China. Clin Microbiol Infect. 2020.

35. Li X, Xu S, Yu M, Wang K, Tao Y, Zhou Y, et al. Risk factors for severity and mortality in adult COVID-19 inpatients in Wuhan. J Allergy Clin Immunol. 2020.

36. Li R, Tian J, Yang F, Lv L, Yu J, Sun G, et al. Clinical characteristics of 225 patients with COVID-19 in a tertiary Hospital near Wuhan, China. J Clin Virol. 2020;127:104363.

37. Zhang JJ, Dong X, Cao YY, Yuan YD, Yang YB, Yan YQ, et al. Clinical characteristics of 140 patients infected with SARS-CoV-2 in Wuhan, China. Allergy. 2020.

38. Shi S, Qin M, Shen B, Cai Y, Liu T, Yang F, et al. Association of Cardiac Injury With Mortality in Hospitalized Patients With COVID-19 in Wuhan, China. JAMA Cardiol. 2020.

39. Goyal P, Choi JJ, Pinheiro LC, Schenck EJ, Chen R, Jabri A, et al. Clinical Characteristics of Covid-19 in New York City. N Engl J Med. 2020.

40. Wang X, Fang J, Zhu Y, Chen L, Ding F, Zhou R, et al. Clinical characteristics of non-critically ill patients with novel coronavirus infection (COVID-19) in a Fangcang Hospital. Clin Microbiol Infect. 2020.

41. Cheng Y, Luo R, Wang K, Zhang M, Wang Z, Dong L, et al. Kidney disease is associated with in-hospital death of patients with COVID-19. Kidney Int. 2020;97(5):829–38.

42. Guan WJ, Ni ZY, Hu Y, Liang WH, Ou CQ, He JX, et al. Clinical Characteristics of Coronavirus Disease 2019 in China. N Engl J Med. 2020;382(18):1708–20.

43. Qin C, Zhou L, Hu Z, Zhang S, Yang S, Tao Y, et al. Dysregulation of immune response in patients with COVID-19 in Wuhan, China. Clin Infect Dis. 2020.

44. Wu J, Li W, Shi X, Chen Z, Jiang B, Liu J, et al. Early antiviral treatment contributes to alleviate the severity and improve the prognosis of patients with novel coronavirus disease (COVID-19). J Intern Med. 2020.

45. Zheng F, Tang W, Li H, Huang YX, Xie YL, Zhou ZG. Clinical characteristics of 161 cases of corona virus disease 2019 (COVID-19) in Changsha. Eur Rev Med Pharmacol Sci. 2020;24(6):3404–10.

46. Yang W, Cao Q, Qin L, Wang X, Cheng Z, Pan A, et al. Clinical characteristics and imaging manifestations of the 2019 novel coronavirus disease (COVID-19):A multi-center study in Wenzhou city, Zhejiang, China. J Infect. 2020;80(4):388–93.

47. Fan N, Fan W, Li Z, Shi M, Liang Y. Imaging characteristics of initial chest computed tomography and clinical manifestations of patients with COVID-19 pneumonia. Jpn J Radiol. 2020.

48. Wu C, Chen X, Cai Y, Xia J, Zhou X, Xu S, et al. Risk Factors Associated With Acute Respiratory Distress Syndrome and Death in Patients With Coronavirus Disease 2019 Pneumonia in Wuhan, China. JAMA Intern Med. 2020.

49. Cai Q, Huang D, Yu H, Zhu Z, Xia Z, Su Y, et al. COVID-19: Abnormal liver function tests. Journal of Hepatology.

50. Zhou F, Yu T, Du R, Fan G, Liu Y, Liu Z, et al. Clinical course and risk factors for mortality of adult inpatients with COVID-19 in Wuhan, China: a retrospective cohort study. Lancet. 2020;395(10229):1054–62.

51. Chen T, Wu D, Chen H, Yan W, Yang D, Chen G, et al. Clinical characteristics of 113 deceased patients with coronavirus disease 2019: retrospective study. BMJ. 2020;368:m1091.

52. Wan S, Xiang Y, Fang W, Zheng Y, Li B, Hu Y, et al. Clinical features and treatment of COVID-19 patients in northeast Chongqing. J Med Virol. 2020.

53. Zhang G, Hu C, Luo L, Fang F, Chen Y, Li J, et al. Clinical features and short-term outcomes of 221 patients with COVID-19 in Wuhan, China. J Clin Virol. 2020;127:104364.

54. Dai H, Zhang X, Xia J, Zhang T, Shang Y, Huang R, et al. High-resolution Chest CT Features and Clinical Characteristics of Patients Infected with COVID-19 in Jiangsu, China. Int J Infect Dis. 2020;95:106–12.

55. Colombi D, Bodini FC, Petrini M, Maffi G, Morelli N, Milanese G, et al. Well-aerated Lung on Admitting Chest CT to Predict Adverse Outcome in COVID-19 Pneumonia. Radiology. 2020:201433.

56. Bai HX, Hsieh B, Xiong Z, Halsey K, Choi JW, Tran TML, et al. Performance of radiologists in differentiating COVID-19 from viral pneumonia on chest CT. Radiology. 2020:200823.

57. Bernheim A, Mei X, Huang M, Yang Y, Fayad ZA, Zhang N, et al. Chest CT Findings in Coronavirus Disease-19 (COVID-19): Relationship to Duration of Infection. Radiology. 2020:200463.

58. Mo P, Xing Y, Xiao Y, Deng L, Zhao Q, Wang H, et al. Clinical characteristics of refractory COVID-19 pneumonia in Wuhan, China. Clin Infect Dis. 2020.

59. Ding X, Xu J, Zhou J, Long Q. Chest CT findings of COVID-19 pneumonia by duration of symptoms. Eur J Radiol. 2020;127:109009.

60. Chen J, Qi T, Liu L, Ling Y, Qian Z, Li T, et al. Clinical progression of patients with COVID-19 in Shanghai, China. J Infect. 2020;80(5):e1–e6.

61. Myers LC, Parodi SM, Escobar GJ, Liu VX. Characteristics of Hospitalized Adults With COVID-19 in an Integrated Health Care System in California. JAMA. 2020.

62. Team CC-R. Coronavirus Disease 2019 in Children - United States, February 12-April 2, 2020. MMWR Morb Mortal Wkly Rep. 2020;69(14):422–6.

63. Liu Y, Du X, Chen J, Jin Y, Peng L, Wang HHX, et al. Neutrophil-to-lymphocyte ratio as an independent risk factor for mortality in hospitalized patients with COVID-19. J Infect. 2020.

64. Grasselli G, Zangrillo A, Zanella A, Antonelli M, Cabrini L, Castelli A, et al. Baseline Characteristics and Outcomes of 1591 Patients Infected With SARS-CoV-2 Admitted to ICUs of the Lombardy Region, Italy. JAMA. 2020.

65. Yin S, Huang M, Li D, Tang N. Difference of coagulation features between severe pneumonia induced by SARS-CoV2 and non-SARS-CoV2. J Thromb Thrombolysis. 2020.

66. Liang W-H, Guan W-J, Li C-C, Li Y-M, Liang H-R, Zhao Y, et al. Clinical characteristics and outcomes of hospitalised patients with COVID-19 treated in Hubei (epicenter) and outside Hubei (non-epicenter): A Nationwide Analysis of China. Eur Respir J. 2020:2000562.

67. Deng Y, Liu W, Liu K, Fang YY, Shang J, Zhou L, et al. Clinical characteristics of fatal and recovered cases of coronavirus disease 2019 (COVID-19) in Wuhan, China: a retrospective study. Chin Med J (Engl). 2020.

68. Guo W, Li M, Dong Y, Zhou H, Zhang Z, Tian C, et al. Diabetes is a risk factor for the progression and prognosis of COVID-19. Diabetes Metab Res Rev. 2020:e3319.

69. Fan Z, Chen L, Li J, Cheng X, Jingmao Y, Tian C, et al. Clinical Features of COVID-19-Related Liver Damage. Clin Gastroenterol Hepatol. 2020.

70. Emerging ISARa, (ISARIC) IC. COVID-19 Report: 08 June 2020. 2020.

71. Jose RJ, Manuel A. COVID-19 cytokine storm: the interplay between inflammation and coagulation. The Lancet Respiratory Medicine. 2020;8(6):e46–e7.

72. Dong Y, Mo X, Hu Y, Qi X, Jiang F, Jiang Z, et al. Epidemiology of COVID-19 Among Children in China. Pediatrics. 2020.

73. Riphagen S, Gomez X, Gonzalez-Martinez C, Wilkinson N, Theocharis P. Hyperinflammatory shock in children during COVID-19 pandemic. The Lancet. 2020;395(10237):1607–8.

74. Pareek M, Bangash MN, Pareek N, Pan D, Sze S, Minhas JS, et al. Ethnicity and COVID-19: an urgent public health research priority. The Lancet. 2020;395(10234):1421–2.

75. Ravi K. Ethnic disparities in COVID-19 mortality: are comorbidities to blame? The Lancet.

76. Bénézit F, Le Turnier P, Declerck C, Paillé C, Revest M, Dubée V, et al. Utility of hyposmia and hypogeusia for the diagnosis of COVID-19. The Lancet Infectious Diseases.

77. Statement from the UK Chief Medical Officers on an update to coronavirus symptoms: 18 May 2020 [press release]. 18/05/2020 2020.

78. Prevention CfDCa. Coronavirus Disease 2019 (COVID-19) 2020 Interim Case Definition, Approved April 5, 2020 Atlanta, Georgia: U.S Department of Health and Human Services; 2020 [Available from: https://wwwn.cdc.gov/nndss/conditions/coronavirus-disease-2019-covid-19/case-definition/2020/.

79. Health AGDo. What you need to know about coronavirus (COVID-19) 2020 [Available from: https://www.health.gov.au/news/health-alerts/novel-coronavirus-2019-ncov-health-alert/what-you-need-to-know-about-coronavirus-covid-19.

80. Bowles L, Platton S, Yartey N, Dave M, Lee K, Hart DP, et al. Lupus Anticoagulant and Abnormal Coagulation Tests in Patients with Covid-19. New England Journal of Medicine. 2020.

81. Zhang Y, Xiao M, Zhang S, Xia P, Cao W, Jiang W, et al. Coagulopathy and Antiphospholipid Antibodies in Patients with Covid-19. New England Journal of Medicine. 2020;382(17):e38.

82. (ISARIC) IASRaeIC. COVID-19 Core Case Report Form Acute Respiratory Infection Clinical Characterisation Data Tool. University of Oxford; 2020.

